# A Direct Capture Method for Purification and Detection of Viral Nucleic Acid Enables Epidemiological Surveillance of SARS-CoV-2

**DOI:** 10.1101/2021.05.06.21256753

**Authors:** Subhanjan Mondal, Nathan Feirer, Michael Brockman, Melanie A. Preston, Sarah J. Teter, Dongping Ma, Said A. Goueli, Sameer Moorji, Brigitta Saul, James J. Cali

## Abstract

Studies have demonstrated that SARS-CoV-2 RNA can be detected in the feces of infected individuals. This finding spurred investigation into using wastewater-based epidemiology (WBE) to monitor SARS-CoV-2 RNA and track the appearance and spread of COVID-19 in communities. SARS-CoV-2 is present at low levels in wastewater, making sample concentration a prerequisite for sensitive detection and utility in WBE. Whereas common methods for isolating viral genetic material are biased toward intact virus isolation, it is likely that a relatively low percentage of the total SARS-CoV-2 RNA genome in wastewater is contained within intact virions. Therefore, we hypothesized that a direct unbiased total nucleic acid extraction method could overcome the cumbersome protocols, variability and low recovery rates associated with the former methods. This led to development of a simple, rapid, and modular alternative to existing purification methods. In an initial concentration step, chaotropic agents are added to raw sewage allowing binding of nucleic acid from free nucleoprotein complexes, partially intact, and intact virions to a silica matrix. The eluted nucleic acid is then purified using manual or semi-automated methods. RT-qPCR enzyme mixes were formulated that demonstrate substantial inhibitor resistance. In addition, multiplexed probe-based RT-qPCR assays detecting the N1, N2 (nucleocapsid) and E (envelope) gene fragments of SARS-CoV-2 were developed. The RT-qPCR assays also contain primers and probes to detect Pepper Mild Mottle Virus (PMMoV), a fecal indicator RNA virus present in wastewater, and an exogenous control RNA to measure effects of RT-qPCR inhibitors. Using this workflow, we monitored wastewater samples from three wastewater treatment plants (WWTP) in Dane County, Wisconsin. We also successfully sequenced a subset of samples to ensure compatibility with a SARS-CoV-2 amplicon panel and demonstrated the potential for SARS-CoV-2 variant detection. Data obtained here underscore the potential for wastewater surveillance of SARS-CoV-2 and other infectious agents in communities.

## Introduction

In late December of 2019, Chinese health authorities examined a new respiratory virus that caused unexplained cases of severe pneumonia^1^. Subsequent sequencing identified the virus as a member of the Coronavirus family, a group of enveloped RNA viruses that commonly infect birds, mammals, and humans^2^. The novel virus, designated Severe Acute Respiratory Syndrome Coronavirus 2 (SARS-CoV-2), is related to SARS-CoV and MERS-CoV, other respiratory viruses that can lead to fatal illness^3^. SARS-CoV-2 was determined to be the causative agent of the respiratory disease COVID-19^1,4^. COVID-19 quickly evolved into a global pandemic, which at the time of this report has resulted in over 3.1 million deaths worldwide (https://coronavirus.jhu.edu/map.html).

The rapid person-to-person spread of SARS-CoV-2 is in part due to the high infectiousness of viral carriers^5,6^. Pre-symptomatic shedding is thought to drive a significant amount of viral spread, as the highest risk of transmission occurs very early in the course of the disease^5,6^. Asymptomatic carriers of the virus are also infectious and display similar viral loads in the respiratory system, despite exhibiting faster viral clearance^6–9^. As traditional nasal swab testing approaches can be biased towards symptomatic viral carriers^10^, viral surveillance methods that provide a widespread view of community infection are vital for accurate monitoring and control of the ongoing pandemic.

Although infectious SARS-CoV-2 virions are rarely isolated from feces^11^, several reports early in the COVID-19 pandemic demonstrated that SARS-CoV-2 can frequently (50%-70%) be detected in fecal samples from both symptomatic and asymptomatic infected individuals^9,12–15^. Levels of SARS-CoV-2 RNA in feces are not correlated with the presence/absence of gastrointestinal illness or with overall COVID-19 disease severity^12^, as is often the case with respiratory samples. Although SARS-CoV-2 viral load is consistently higher in respiratory specimens, viral RNA in feces can be detected significantly longer after initial symptom onset compared to nasal swab samples^13,16^. This aspect of SARS-CoV-2 fecal shedding could be a useful feature for assaying the levels of overall infection in a community.

One method that has been proposed for use in monitoring population-level rates of SARS-CoV-2 infection is wastewater-based epidemiology (WBE). WBE is the quantitative detection of chemical and biological signatures in wastewater to analyze the status of a human population in an area^17^. This approach has been used extensively in the past to characterize community usage/prevalence of illicit drugs such as cocaine, amphetamines, and opiates^17^. WBE detection methods have also been widely used to monitor the trends of circulating human pathogens in wastewater streams around the world^17,18^. Of particular relevance to SARS-CoV-2 are the efforts centering around environmental surveillance of the enteric viruses Poliovirus, Hepatitis A, and Norovirus^19–21^

In the Spring of 2020, research groups around the world started reporting the detection of SARS-CoV-2 RNA in untreated wastewater^22–25^ using quantitative reverse transcription PCR (RT-qPCR). SARS-CoV-2 RNA levels in wastewater positively correlated with clinical COVID-19 case numbers^22^. Several studies were able to demonstrate that an increasing trend of SARS-CoV-2 in wastewater preceded a rise in clinical cases by up to one week^25–27^, establishing the potential utility of COVID-19 WBE as an “early warning system” to mitigate viral spread and cost effective strategy for monitoring real-time changes in viral prevalence.

As viral concentrations in wastewater tend to be low, sample concentration is often a pre-requisite for sensitive detection and accurate quantitation. Concentration of viral matter can be performed using a variety of methods such as charged membrane filtration, centrifugal ultrafiltration, flocculation/precipitation using skimmed milk, and polyethylene glycol (PEG)/NaCl precipitation^28^. Most of the viral concentration methods described in the literature were originally developed to concentrate non-enveloped viral particles for use in downstream culture-based approaches, but they have been also used for PCR detection. Many of these methods may work efficiently with several virus types but present difficulties when applied to SARS-CoV-2. Notable issues have included inconsistent rates of viral recovery after concentration, requirement for large sample sizes, and co-purification of PCR inhibitors^29,30^. In addition, these viral concentration methods are labor intensive and time consuming, requiring separate viral concentration and a nucleic acid extraction steps.

In this report, we describe the use of a novel column-based viral concentration and nucleic acid purification system coupled to RT-qPCR detection for monitoring SARS-CoV-2 levels in wastewater. The system allows for increased sample throughput due to the combination of viral purification and nucleic acid purification steps. The effect of PCR inhibitors commonly found in wastewater is also minimized. To demonstrate proof-of-concept, wastewater from three communities in Dane County, Wisconsin was monitored for the presence of SARS-CoV-2 RNA over the course of three months. We also successfully sequenced from a subset of samples to show compatibility with a commercially available SARS-CoV-2 amplicon panel and demonstrated the potential for SARS-CoV-2 variant detection from wastewater nucleic acid purified with the direct capture method.

## Methods

### Sample collection

Wastewater (untreated primary effluent) was collected from three wastewater treatment plants in Dane County, Wisconsin: Oregon (WWTP-1), Madison (WWTP-2), Sun Prairie (WWTP-3). In all three communities, the wastewater collection system is separate from the storm sewer system, minimizing the dilution effect of precipitation events. 500-1000mL of a flow-paced (Oregon, Sun Prairie) or time-paced (Madison) 24-hour composite sample were collected weekly using an autosampler (Madison: ISCO FR3710; Sun Prairie: ISCO 5800; Oregon: ISCO 4700). Samples were kept at 4°C at all times during transport and storage. All samples were processed within six hours of sampling.

MS2 (ATCC 15597-B1) and *Escherichia coli* ATCC 15597 were purchased from American Type Culture Collection (ATCC) and propagated based on published methods^31^. The viral fraction in the culture supernatant was collected by centrifuging at 3500 x *g* for 30 minutes to remove the bacterial fraction and filtered through a 0.22 μm PES membrane filter. The purified MS2 virus (∼10^7^ PFU mL^−1^) was stored at 4 °C. MS2 virus was spiked in wastewater samples to final concentrations of 4 × 10^4^ PFU mL^−1^. Samples were then briefly mixed and nucleic acid extracted as described below.

### Isolation of Total Nucleic Acid from wastewater using the direct capture method

Total nucleic acid (TNA) was purified from collected wastewater using the Wizard^®^ Enviro Wastewater TNA kit (Promega Corp.) and/or the Maxwell^®^ Enviro Wastewater TNA kit (Promega Corp.), both of which use an initial concentration step composed of direct capture of nucleic acids on silica resin. Briefly, 0.5 mL of alkaline protease was added to 40 mL of untreated wastewater in triplicate, and samples were incubated statically for 30 minutes at room temperature. The samples were then centrifuged at 3000 x *g* for 10 minutes to remove suspended solids. The supernatant was transferred to a new vessel and 12 mL of Binding Buffer 1 and 1 mL of Binding Buffer 2 were added, followed by gentle mixing. A 48 mL of isopropanol was added to the mixture, gently mixed and passed through a PureYield™ Midi Binding Column (Promega Corp.) using a VacMan^®^ Vacuum Manifold (Promega Corp.). Nucleic acid captured on the PureYield™ Midi Binding Column was washed with 5 mL of Column Wash 1 followed by 20 mL of Column Wash 2. Nucleic acid was eluted with nuclease-free water (Fig. 2).

The eluted nucleic acid was further purified using a Mini spin column for the Wizard^®^ Enviro Wastewater TNA kit or with an automated nucleic acid purification system (Maxwell® RSC, Promega Corp) for the Maxwell^®^ Enviro Wastewater TNA kit (Fig. 2). For the manual Wizard^®^ Enviro Wastewater TNA kit, 400 µL of Binding Buffer 1, 100 µL of Binding Buffer 2 and 1.5 mL of isopropanol are added to the 1 mL of nucleic acid extracted in the concentration step, and then passed through a spin column with a silica resin. The column is washed with 350µL and 1 ml of Column Wash 1 and 2 respectively, and nucleic acid is extracted in 80 µL of water. For the automated Maxwell^®^ Enviro Wastewater TNA kit, 150 µL of Binding Buffer 1 and 50 µL of Binding Buffer 2 are added to 0.5 mL of nucleic acid extracted in the concentration step. Then the total volume is added to well #1 of the Maxwell^®^ Cartridge and nucleic acid is eluted in 80 µL of nuclease-free water.

### Isolation of Total Nucleic Acid from suspended solids using the direct capture method

The method to extract total nucleic acids from suspended solids is a variation on the direct capture methodology described above. After the alkaline protease step, the pellet of suspended solids is resuspended in 5 mL of nuclease-free water. To the resuspended pellet, 1.5 mL of Binding Buffer 1, 125 μL of Binding Buffer 2 and 6 mL of isopropanol were added and mixed. This step releases the nucleic acid bound to the solids into the suspension. The mixture was centrifuged at 3,000 x *g* for 10 minutes. The supernatant contains the nucleic acid from the solids. The supernatant is then added to the PureYield™ Midi Binding Column for the respective sample and treated independently as described above.

### PEG/NaCl precipitation

A sample of 120 mL of wastewater was centrifuged at 3000 x *g* for 30 minutes to pellet any particulate material. The supernatant was carefully decanted and then mixed with 12 g of PEG 8000 and 2.7 g of NaCl that were dissolved by gentle mixing. Samples were then centrifuged at 11,400 x *g* for two hours to pellet viral material. The supernatant was carefully removed via pipetting on the side of the tube opposite of that which the pellet was formed (pellet was not visible). The pellets were resuspended by vortexing with residual supernatant to a final volume of approximately 200 µL. Further nucleic acid purification was performed using Maxwell^®^ RSC PureFood GMO and Authentication Kit (Promega Corp.) using the Maxwell^®^ RSC Instrument (Promega Corp.) according to a modified version of manufacturer’s protocol. Briefly, to 200μl of wastewater concentrate, 200 μL of CTAB and 40 μL of Proteinase K were added. Samples were vortexed, then incubated at 56°C for 10 minutes. The entire sample volume and 300 μL of Lysis Buffer were added to well #1 of the Maxwell^®^ RSC cartridge. Nucleic acid was eluted with 50 μL of Elution Buffer. Samples were processed on the Maxwell^®^ RSC with the PureFood GMO and Authentication method.

### Oligonucleotides and quantification standards

Table S1 lists the primers and probes used in this study, including primer and probe sets for the detection of SARS-CoV-2 (CDC-N1, CDC-N2, E_Sarbeco), PMMoV, and MS2 bacteriophage. The primers and probe for amplifying the luciferase (*luc)* gene for the internal amplification control are also listed.

The SARS-CoV-2 quantification standard was created by inserting the envelope gene (NCBI: 43740570) and the nucleocapsid gene (NCBI: 43740575) of SARS-CoV-2 into the pGEM-3z vector (Promega Corp.) using the *Bam*H I site. The plasmid was subsequently linearized using *Xba* I. The linearized plasmid was in-vitro transcribed using T7 RiboMAX™ Large Scale RNA Production System (Promega Corp.) to create Positive Control RNA. The linearized plasmid DNA containing the N and E genes, or the in-vitro transcribed RNA were quantified using droplet digital PCR and used as a quantification control. Linear quantification DNA and positive control RNA for PMMoV and MS2 were generated and quantified using the same methods described above.

### RT-qPCR and quantification of viral load in wastewater

TNA isolated from wastewater was used to perform RT-qPCR using the SARS-CoV-2 RT-qPCR Detection Kit for Wastewater (Promega Corp.). RT-qPCR reactions targeted the nucleocapsid (N1, N2) and envelope (E) regions of the SARS-CoV-2 genome. To allow for quantitation, log dilutions (2×10^1^ – 2×10^5^ Genome Units (GU)/µL) of DNA quantitation standard were amplified alongside experimental samples and used to construct a standard curve for each SARS-CoV-2 target. 20 µL amplification reactions were composed of 15 µL reaction mastermix and 5uL of nucleic acid. 5 µL of nuclease-free water was used as a no-template-control (NTC). Reactions were performed on a Stratagene MX3005 Real-Time Thermocycler (Agilent) with the following cycling conditions: reverse transcription for 15 minutes at 45°C, initial denaturation for 2 minutes at 95°C, and 40 cycles of 3 seconds at 95°C and 30 seconds at 62°C. Standard curve reactions were run in triplicate and wastewater sample reactions were run in duplicate. The standard curve approach described above was used in parallel to estimate the concentration of MS2 and PMMoV in purified samples.

Concentration of viral load in wastewater was calculated using the equation:

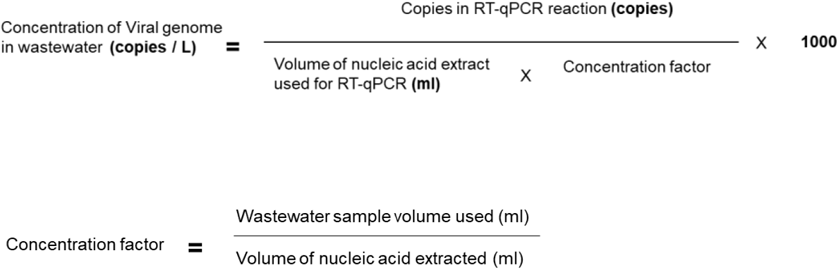

Normalization of SARS CoV-2 amounts in wastewater was performed by using the following equation:

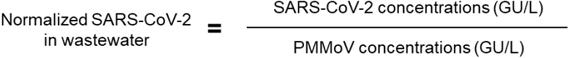

### Statistical analysis

Concordance between trend analysis using the three SARS-CoV-2 targets (N1, N2 and E) was evaluated using Kendall’s coefficient of concordance (W). Correlation between weekly normalized SARS-CoV-2 levels from wastewater and number of new clinical cases (7 day moving average) was determined using Kendall’s tau correlation coefficient.

### SARS-CoV-2 amplicon library preparation and sequencing

10 µL of each total nucleic acid sample was treated for 30 minutes at 37°C with 1.5 U of RQ1 RNase-Free DNase (Promega Corporation) in a 15 µL reaction, as specified in the manufacturer’s protocol, except that the Stop Solution provided was not used. 10 µL of nuclease-free water was added to each sample and cleaned up with the ReliaPrep™ RNA Clean-Up and Concentration System (Promega Corporation), as specified in the manufacturer’s technical manual. RNA was eluted with 15 µL of nuclease-free water.

cDNA was synthesized as follows. 10 µL of either total nucleic acid sample or DNase-treated sample was used as a template for first strand synthesis with random hexamers and associated steps using Invitrogen™ SuperScript™ IV First-Strand Synthesis System (Thermo Fisher Scientific). Manufacturer’s instructions were followed with one exception: incubation time at 50°C was increased from 10 minutes to 30 minutes, as recommended by the Swift Biosciences SARS-CoV-2 Additional Genome Coverage amplicon panel library preparation protocol.

10 µL of cDNA was used as input for the SARS-CoV-2 Additional Genome Coverage panel (Swift Biosciences) using the low input version of the library preparation protocol. This 345-amplicon panel covers 99.7% of the SARS-CoV-2 genome and has amplicons ranging from 116 to 255 bp (average 150 bp). Libraries were quantified by qPCR, pooled, and sequenced with 2 × 150 base-pair reads on an Illumina MiniSeq Instrument with a MiniSeq Mid Output Kit (300-cycles).

### Amplicon sequencing analysis

Compressed, demultiplexed reads were obtained from the Illumina MiniSeq instrument and assessed for sequencing quality using FastQC 0.11.9-0 (https://www.bioinformatics.babraham.ac.uk/projects/fastqc/). For the purpose of comparing performance between libraries, random subsampling was performed to obtain the same number of input reads for each library using seqtk v1.2. All available reads for each library were used in known variant detection. Adapter trimming was done using Fastp v0.20.1^32^ (sequences AGATCGGAAGAGCACACGTCTGAACTCCAGTCA and AGATCGGAAGAGCGTCGTGTAGGGAAAGAGTGT). Trimmed reads were mapped to the SARS-CoV-2 reference genome (NC_045512.2) using BWA v 0.7.17. To assess human genomic contribution, trimmed reads were also mapped to a combined SARS-CoV-2 + GRCh38/hg38^33^ (UCSC Genome Browser^34^ Dec 2013 assembly) reference genome. Amplicon primers were trimmed following alignment using Swift’s Primerclip tool v 0.3.8 (https://github.com/swiftbiosciences/primerclip) with the Swift-provided master file for the 345 amplicon panel as input. QC metrics were generated using Picard (v 2.9.2; http://broadinstitute.github.io/picard) BedToIntervalList, CollectTargetedPcrMetrics, CollectGcBiasMetrics. The GATK3^35^ DepthOfCoverage tool was used for target base coverage assessments (median, mean, percent of bases at or above 1X, 100X, 1000X, and 5000X). Snakemake v5.17.0 was used for workflow management on a Microsoft Azure CycleCloud instance.

### Detection of known signature variants

Genomic locations of known signature variants were obtained from Nextstrain^36^ (accessed 12 March 2021) and UCSC Table Browser^37^ (SARS-CoV-2 Jan. 2020/NC_045512.2 Assembly (wuhCor1)) and formatted into a BED file. With aligned, primer-trimmed reads in BAM format, the bam-readcount tool (https://github.com/genome/bam-readcount) was used to collect base composition information at the signature variant locations. The bam-readcount output was filtered to look for the presence of signature variants with at least 10% frequency at positions covered to at least 50X, so that no signature variant would be called as detected with fewer than 5 sequencing reads as evidence.

## Results

### RT-qPCR Reaction Formulation and Setup

Operating under the assumption that wastewater is likely to contain RT-qPCR inhibitors, we formulated enzyme mixes containing MMLV-RT enzyme and Taq DNA polymerase (with hot-start chemistry) that would be resistant to PCR inhibitors. Next, we designed separate multiplexed RT-qPCR assays to detect the nucleocapsid gene fragment N1 and N2 (as described by the CDC: https://www.cdc.gov/coronavirus/2019-ncov/lab/rt-pcr-panel-primer-probes.html) and the envelope gene (as described by Corman *et al*.^38^). SARS-CoV-2 targets were detected using primers and a target-specific hydrolysis probes (labeled with 5’ FAM/ZEN™/ 3’ Iowa Black™ FQ). Each assay was multiplexed with primers and a hydrolysis probe (labeled with 5’ Quasar^®^670/ BHQ^®^-2) complementary for Pepper Mild Mottle Virus (PMMoV). PMMoV is a single-stranded RNA plant virus that commonly infects pepper products intended for human consumption^39^. PMMoV RNA is detectable in wastewater worldwide and is considered an important indicator of human-derived fecal pollution^40–42^. Each assay also includes an exogenous RNA template, primers and hydrolysis probe (labeled with 5’ CalFluor^®^ Orange 560/ BHQ^®^-1) comprising an internal amplification control (IAC). The IAC’s cycle threshold (Ct) provides information on the presence of reverse transcriptase and DNA polymerase inhibitors in the extracted nucleic acid sample. CXR, (Carboxy-X-Rhodamine, Promega Corp.) which has similar spectral properties as ROX (Ex: 580nm, Em: 602nm) was used as a reference dye.

We sought to optimize an appropriate amplicon length from the IAC template to provide sufficient sensitivity to detect RT-qPCR inhibition. An *in vitro* transcribed Luciferase RNA (Promega Corp.) was used as a template and four different amplicon lengths (93 bp, 285 bp, 310 bp and 435bp) were tested. Humic acid, a known reverse transcriptase and DNA polymerase inhibitor^43^, was titrated from 0 µg/mL to 125 µg/mL using a two-fold dilution series, and inhibition was assessed by the difference in Ct value with or without humic acid (ΔCt). The same hydrolysis probe was used in all cases. As expected, we found a correlation between the amplicon length and sensitivity to humic acid (Fig. 1A), with the 435 bp and 315 bp amplicons exhibiting higher ΔCt values compared to the 285 bp and 93 bp amplicons. Since the RT-qPCR amplicons for detection of SARS-CoV-2 and PMMoV are below 150 bp in length and the ΔCt values for 285bp and 93bp amplicons were similar, the 285 bp amplicon length was used as IAC unless otherwise specified.

**Figure 1.**
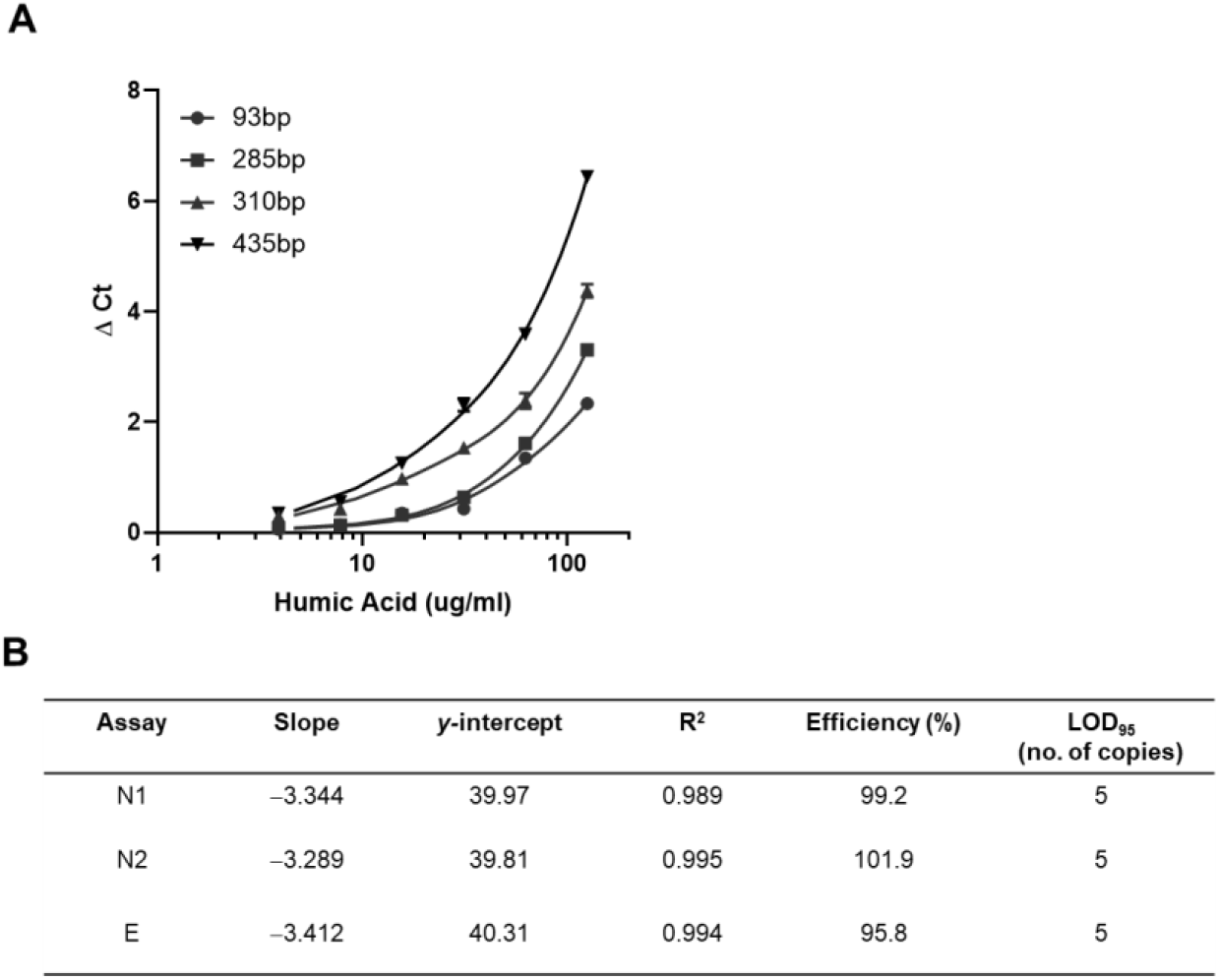
(A) Optimization of Internal Amplification Control (IAC) amplicon size for indication of inhibitor tolerance. Amplification of *luc* in-vitro transcribed RNA template of listed amplicon sizes was performed with different humic acid concentrations present in the RT-qPCR reaction. (B) Performance characteristics of the three multiplexed RT-qPCR assays for detection of SARS-CoV-2 in wastewater. Standard curve reactions were performed in triplicate.

For WBE, it is important to quantify the viral genome units per volume of wastewater to determine the quantitative trend in viral load. To analyze the analytical sensitivity, efficiency, and linearity of the assay, a log dilution series of the in-vitro transcribed SARS-CoV-2 RNA (N and E) was used to perform RT-qPCR analysis. PCR amplification efficiencies for all three targets were between 90-120%. The limit of detection (LOD_95_) for the three multiplexed assays for detecting the SARS-CoV-2 targets was 5 copies and a limit of quantification (LOQ) of 8 copies. The RT-qPCR assays are linear in the tested range of 20-200,0000 copies, as the observed R^2^ for all the three targets were ≥ 0.99 (Fig. 1B).

The assay was also tested for specificity with other coronaviruses and respiratory pathogens. The assay was found to be specific for detection of SARS-CoV-2 and PMMoV (Table S2).

### Description of the direct-nucleic acid capture method

Existing literature suggests that SARS-CoV-2 may not be infectious in wastewater samples^44,45^. It is therefore unknown if the viral genetic signature present in the wastewater samples is entirely derived from compromised virions or if some proportion is present as unpackaged SARS-CoV-2 nucleic acid. We hypothesized that by utilizing a direct capture method to bind total nucleic acid (TNA) to a silica-based affinity resin in place of a method that is selective for intact viral particles, we may be able to eliminate the viral concentration step which is often a cause of technical variability and low recovery. Direct TNA isolation would be unbiased toward intact, partially intact, or free viral RNA. We developed a simple, rapid, highly efficient, and modular alternative to existing wastewater RNA purification methods. The primary concentration method utilizes raw sewage, to which chaotropic agents are added to allow binding on to a silica matrix (PureYield™ Midi Binding column) by applying vacuum. The captured nucleic acid is then subjected to successive alcohol washes (to remove RT-qPCR inhibitors that may have co-purified with the nucleic acid) and subsequently eluted in water (Fig. 2). The eluted nucleic acid can then be further processed in a second step with either a spin column with Wizard^®^ Enviro Wastewater TNA kit or with an automated nucleic acid purification system for the Maxwell^®^ RSC Enviro Wastewater TNA kit (Fig. 2).

**Figure 2.**
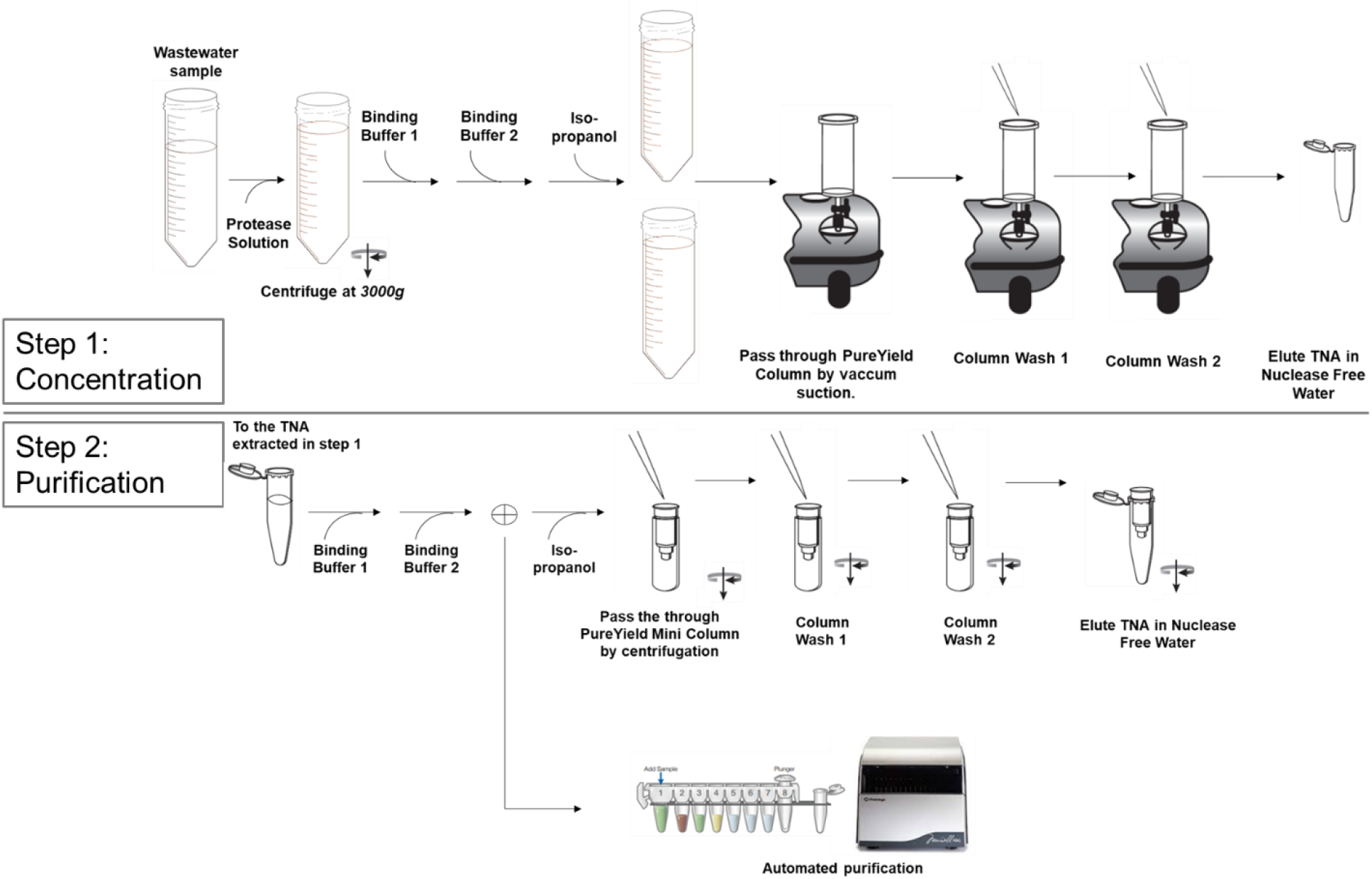
Schematic overview of the direct capture nucleic acid purification process.

The direct-nucleic acid capture method was first tested for its ability to eliminate RT-qPCR inhibitors. Inhibition was analyzed by comparing the difference in Ct value (ΔCt) between IAC amplification in reactions for wastewater sample and for no-template-control (NTC) reactions. The ΔCt values were analyzed for samples processed using either the primary concentration step with the PureYield™ Midi column only or processed through the complete workflow comprising the Wizard^®^ concentration/purification steps outlined above. ΔCt values > 1 indicated the presence of reverse transcriptase and/or DNA polymerase inhibitors in the wastewater samples added to the RT-qPCR reaction. ΔCt values of <1 were observed for both nucleic acid purified with the PureYield™ Midi column only and samples purified with both ™ Midi and PureYield™ Mini column steps (Fig. 3A). In the second step, total nucleic acid eluted from the first step is further purified in a smaller volume (80 µL), concentrating the nucleic acid by 12.5-fold. This concentration is evident when MS2 (viral spike-in control) or SARS-CoV-2 (N1) is analyzed, as we observed a 8.34- and 5.54-fold increase in the amount of MS2 and SARS-CoV-2 RNA detected, relative to the sample that only underwent initial sample concentration (Fig. 3B and C). This indicates that the second purification step successfully concentrates the nucleic acid in the sample.

**Figure 3.**
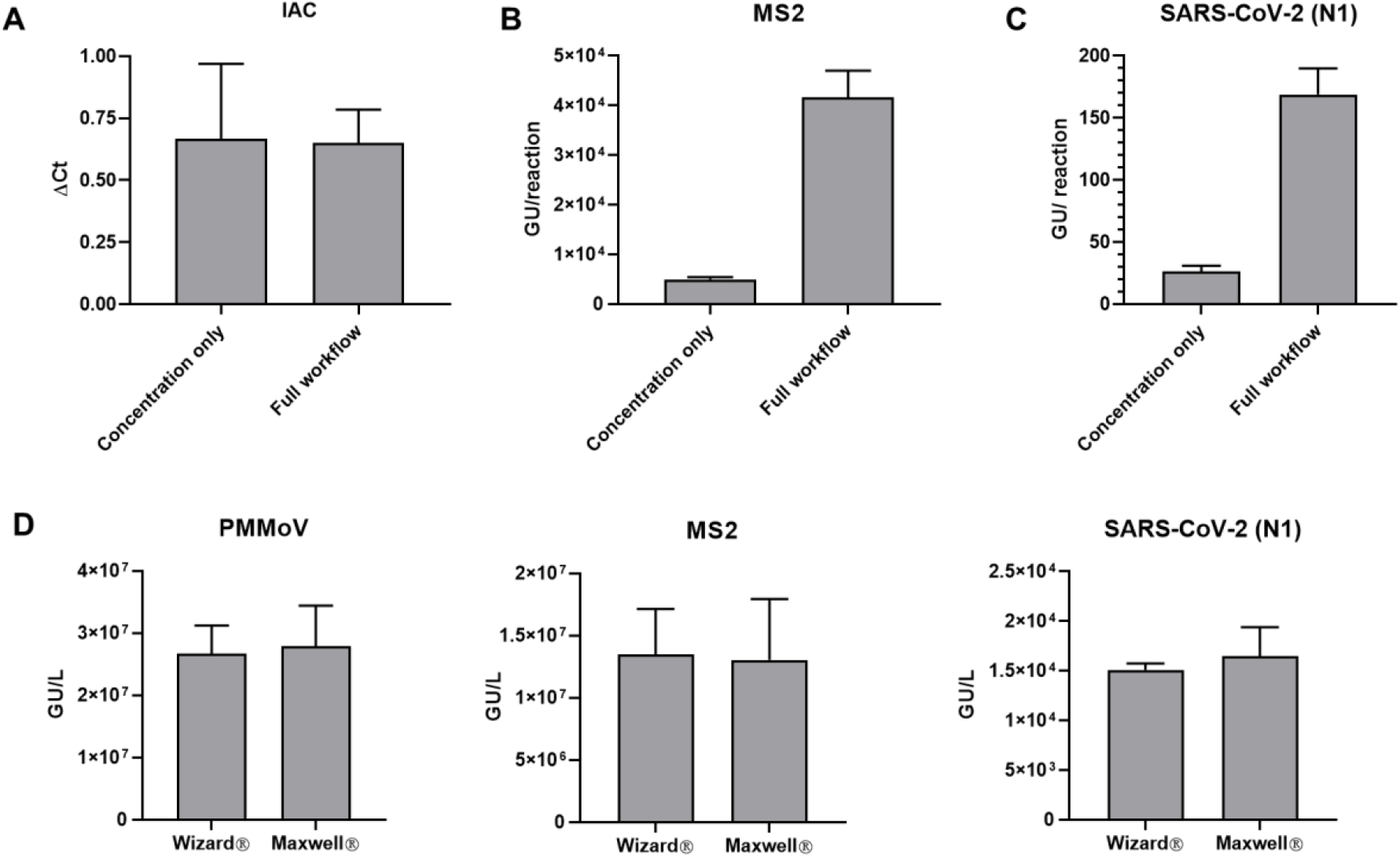
(A) The direct capture method can reduce presence of RT-qPCR inhibitors present in extracted samples. “Full workflow” refers to the combination of concentration and purification steps shown in Fig. 2. ΔCt is defined as the difference in internal amplification control (IAC) amplification (Ct Value) between the NTC wells compared to sample wells. Viral concentrations detected via RT-qPCR using listed workflows for (B) MS2 phage (viral wastewater spike) and (C) SARS CoV-2. (D) The extraction efficiencies of the purification step performed using the Wizard spin column or Maxwell automated instrument are similar for the extraction of PMMoV, MS2 and SARS-CoV-2. Results shown are means ± SD (*n=* 3)

Different laboratories have different throughput needs. Because our workflow can be done manually or using automation for the final sample concentration, it provides flexibility for different types of labs. This flexibility allows scale-up of the nucleic acid concentration process using an automated nucleic acid purification workflow after the initial concentration step using the PureYield™ Midi column. We compared the extraction efficiency of the nucleic acid isolated by either the manual spin column (Wizard^®^ Enviro Wastewater TNA kit) or with an automated nucleic acid purification system for the Maxwell^®^ RSC Enviro Wastewater TNA kit. Purified concentrations of PMMoV, MS2 (viral spike), and SARS-CoV-2 RNA were similar with both purification methods (Fig. 3D). These data indicate that both manual and automated nucleic acid purification procedures can be used to extract viral genetic material at similar extraction efficiencies.

### Sample volume considerations

SARS-CoV-2 levels in wastewater are often very low, making sample concentration from a larger volume a necessary part of any WBE monitoring workflow. To determine what starting sample volume allows for convenient and sensitive viral detection, we processed different volumes of wastewater (80 mL, 40 mL, 20 mL, 10 mL, and 5 mL) using the column-based manual concentration/purification scheme (Wizard^®^) outlined above and performed all final elutions in the same volume (80 µL) of nuclease-free water. The adjusted volumes of Protease solution, binding buffers 1 and 2 and isopropanol used for each starting volume are outlined in Fig. 4A. MS2 bacteriophage was used as a viral spike-in recovery control.

**Figure 4.**
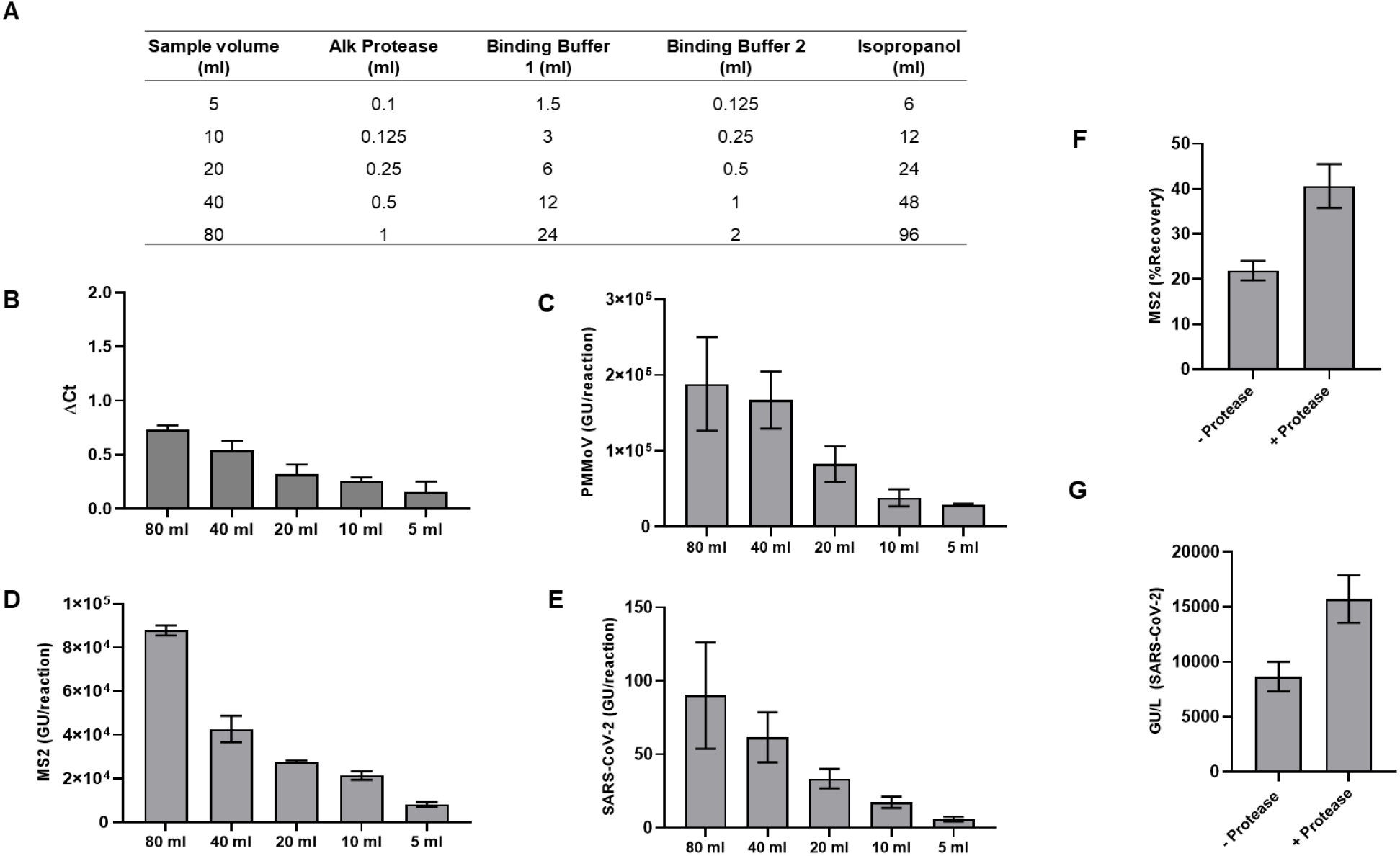
Sample volume considerations. (A) Volumes of reagents required for different sample volumes. (B) Increasing the sample volume from 5 ml to 80 ml does not lead to significant inhibition of RT-qPCR performance as measured using the Internal amplification control (IAC). DCt represents the difference in Ct values of samples compared to the NTC reactions. (C-E) Increase in the amount of viral genetic material extracted as sample volume is increased as measured by RT-qPCR detection of PMMoV (C), MS2 (D) and SARS-CoV-2 (E). Extraction efficiency of MS2 (F) and SARS-CoV-2 (G) genetic material from wastewater samples with and without protease treatment. Results shown are means ± SD (*n=* 3).

The shift in Ct values from the NTC reactions (ΔCt) for the IAC exhibited a sample volume-dependent increase in Ct value indicating a higher concentration of inhibitors may be co-purifying when larger volumes are used. However, the ΔCt was <1 Ct for all the volumes tested, indicating co-purification of RT-PCR inhibitors was not high enough to significantly impact data interpretation (Fig 4B). We also analyzed the amounts of PMMoV, MS2 and SARS-CoV-2 (N1). As expected, all targets showed a volume-dependent enrichment of genetic material. (Fig 4C-E). Though we can accurately detect SARS-CoV-2 signal from 5 ml of sample during the current sampling period when SARS-CoV-2 clinical cases are high, use of 40 ml sample volume will allow sufficient assay sensitivity when viral loads are lower.

### Enhancement of viral nucleic acid recovery with protease treatment

Wastewater, in addition to containing fecal matter and water, is also composed of cellulosic material from toilet and tissue paper that may act as a substrate on which nucleic acids and nucleoprotein complexes can aggregate. This material may form a large part of the suspended solids present in wastewater. SARS-CoV-2 may not be present as intact virions in wastewater, but the observation that the genetic material is readily detectable indicates that the viral RNA is likely present in ribonucleoprotein complexes, which shield it from nucleases that are presumably present in wastewater. In addition, detergents and chaotropic agents present in wastewater may also cause structural changes to proteins causing association with suspended solids. Therefore, we reasoned that a proteolytic cleavage step may be able to release some of the viral genetic material associated with solids.

We measured viral nucleic acid extraction efficiency using a procedure that included alkaline protease treatment. 40 mL wastewater samples were either treated with alkaline protease or left untreated and processed as described in the Methods section. MS2 phage was also spiked into the samples. Percent recoveries of MS2 nucleic acid were found to be 20% for untreated samples and 40% for samples that were treated with alkaline protease (Fig. 4F). Similarly, we also observed a two-fold increase in extraction of SARS-CoV-2 viral genome units when samples were treated with protease (Fig. 4G). These results indicate that alkaline protease treatment increases yield of viral TNA extraction, most likely by releasing a portion of the viral genomic material associated with suspended solids.

### Amount of viral matter associated with solids

In the purification workflow described above, suspended solids are removed via a brief centrifugation following the alkaline protease treatment step. Removing suspended solids prevents clogging of the PureYield™ Midi column. We investigated how much viral matter is associated with the pelleted solids using the procedure described in the methods section. The eluted nucleic acid was analyzed for the quantity of PMMoV and SARS-CoV-2 RNA. We observed that about 11.79% of the total amount of PMMoV and 19.27% of the total amount of SARS-CoV-2 genetic material associated with the solids (Fig. 5A-B). If maximal nucleic acid yield is desired, the solids-associated supernatant fraction can be combined with the wastewater supernatant fraction to purify nucleic acid from the total pooled sample.

**Figure 5.**
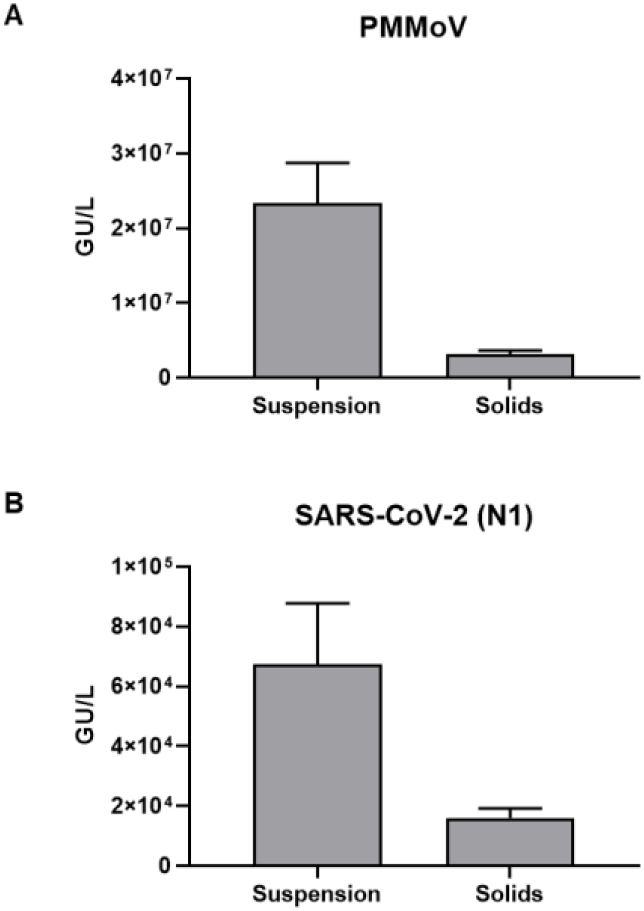
Amount of viral matter associated with solids after protease treatment. Wastewater samples were centrifuged after protease treatment and resulting solids and suspension fractions were processed as described in the *Methods* section. Concentrations of PMMoV (A) and SARS-CoV-2 (B) RNA present in the two fractions as measured by RT-qPCR are shown. Results shown are means ± SD (*n=* 3).

### Comparison of viral nucleic acid recovery: Direct Capture method vs PEG/NaCl precipitation

Using the optimized method described above, we compared the direct capture method with PEG/NaCl precipitation method for the ability to purify SARS-CoV-2 genetic material from wastewater samples. PEG/NaCl is a widely used method for precipitation and concentration of non-enveloped enteric virus such as Poliovirus^28^. 40 mL of wastewater sample was processed for the direct capture method and 120 mL of wastewater was processed using PEG/NaCl precipitation. We observed a 20-fold increased extraction efficiency for extracting SARS-CoV-2 RNA (Fig. 6A) when using the direct capture method compared to the PEG/NaCl protocol. For MS2 (viral spike control) we observed an extraction efficiency of 3.76 ± 1.88% for the PEG/NaCl method and 39.67 ± 10.66% for the direct capture method (Fig 6B). We also determined the percentage recovery for two human coronaviruses (OC43 and 229E) for the direct capture method and they were 63.13 ± 4.16%, 40.09 ± 10.89% respectively (data not shown).

**Figure 6.**
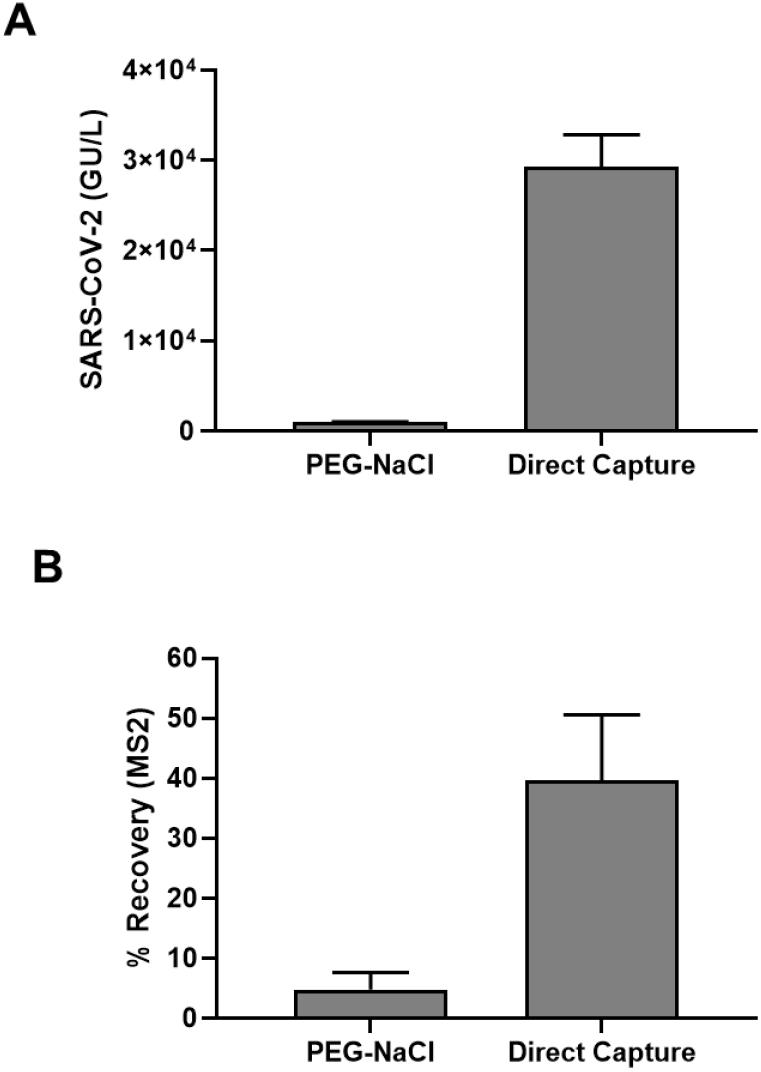
Comparison of the direct capture method with PEG/NaCl precipitation. Wastewater samples were processed using the two methods as described in the *Methods* section. (A) Concentration of SARS-CoV-2 RNA extracted is represented as GU/L in the wastewater sample, (B) Percentage recovery for extraction of MS2 phage (viral spike control). Results shown are means ± SD (*n=* 3).

The direct capture purification workflow utilizing 40 ml of wastewater sample and a 40ul elution volume has a concentration factor of 1000. With a LOD of 5 copies for the detection of SARS-CoV-2 by RT-qPCR reaction, and 1000-fold concentration in the purification process, the assay sensitivity is around 1 viral genome copies/ml. This level of sensitivity will be sufficient for trend analysis using WBE.

### SARS-CoV-2 RNA in wastewater from Dane County, Wisconsin

The levels of SARS-CoV-2 RNA from three wastewater treatment plants in Dane County, Wisconsin from mid-October, 2020 to early January, 2021 were determined using the Wizard^®^ Enviro Wastewater TNA kit described above. Samples were processed and analyzed weekly. The levels of RT-qPCR inhibitors present in the TNA samples (as assessed by shift in the Ct value (ΔCt) of IAC in sample wells compared to NTC wells) were not notable, as ΔCt values were less than 0.5 for all wastewater samples (Fig. 7A).

**Figure 7.**
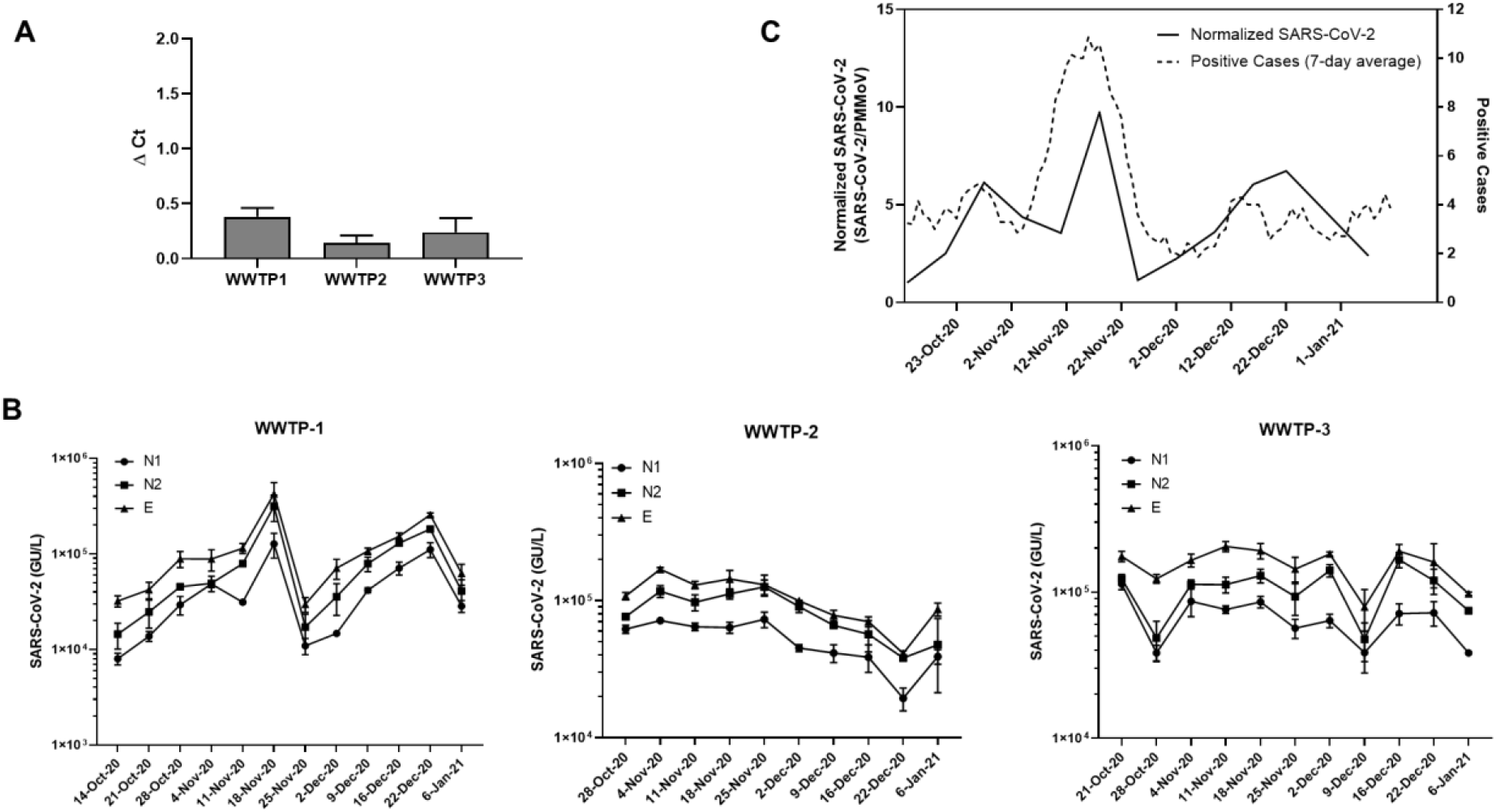
WBE analyses of samples from three wastewater treatment plants in Dane County, WI. (A) Assessment of PCR inhibitors in nucleic acid isolated from the three WWTP. Results shown are means ± SD (*n>* 30). (B) Levels of SARS-CoV-2 analyzed over the indicated duration with three SARS-CoV-2 targets (N1, N2, E) for the three WWTP. Results shown are means ± SD (*n=* 3). (C) Comparison of the normalized SARS-CoV-2 RNA levels with the 7-day moving average of the new reported cases for the community serving WWTP-1.

We analyzed SARS-CoV-2 RNA levels using N1, N2, and E targets (Fig. 7B). All samples were positive for all three SARS-CoV-2 targets within the period tested. We calculated the degree of concordance between the three SARS-CoV-2 targets for the three WWTPs over the sampling period using Kendall’s coefficient of concordance, *W*. Kendall’s *W* ranges from 0 (no agreement) to 1 (full agreement). We observed statistically significant concordance between the three SARS-CoV-2 targets for the three WWTPs (W=0.97 for WWTP-1, W=0.96 for WWTP-2 and W=0.78 for WWTP-3).

WWTP-1 served a small community of around 10,000 people. We normalized SARS-CoV-2 levels with PMMoV, which has been proposed to account for differential dilution and degradation rates over time. We compared SARS-CoV-2 RNA signals in wastewater to the level of COVID-19 cases (7-day moving average) declared by the municipality and analyzed the correlation between the two, resulting in a Kendall’s Tau coefficient of 0.33, with a p-value of 0.08 (Fig 7C). The peak of SARS-CoV-2 genetic signal observed in the wastewater is concurrent with the peak of positive SARS-CoV-2 reported in mid-November of 2020. Even with this limited data set we see the potential for wastewater-based surveillance in assessing community-wide spread of the disease.

### Amplicon sequencing of SARS-CoV-2 genetic material in wastewater

To determine if our direct capture purification method yielded nucleic acid compatible with next-generation sequencing, we prepared sequencing libraries with a subset of wastewater samples using a commercially available SARS-CoV-2 amplicon panel and associated library preparation kit^46,47^.

For this proof of concept, we focused on three samples: wastewater from WWTP-2 in November of 2020 and January 2021, as well as wastewater collected from WWTP-3 in December of 2020. Because of the proximity of the two collection sites to one another, we reasoned that these samples could be compared broadly for the purpose of identifying SARS-CoV-2 variants, while also demonstrating the robustness of the method to different wastewater treatment regimens from different facilities.

We compared total nucleic acid or DNase-treated nucleic acid as input into the library preparation workflow. Most libraries had greater than 1 million reads, but for comparison of depth metrics, the total number of reads was normalized across libraries by randomly subsampling to 650,000 total paired reads. We aligned the subsampled, filtered sequencing reads to the SARS-CoV-2 genome and measured the percentage of filtered reads aligned and the depth of coverage across amplicons tiling the SARS-CoV-2 genome (Fig. 8A-B). Samples that were DNase-treated had higher percentage of filtered reads aligning to the SARS-CoV-2 genome and greater depth of coverage compared to total nucleic acid libraries. Because wastewater is a heterogeneous sample, it is not surprising that the amplification reaction resulted in off-target amplification. This data shows that DNase treatment of wastewater total nucleic acid improves next-generation sequencing quality from the SARS-CoV-2 amplicon panel tested.

**Figure 8.**
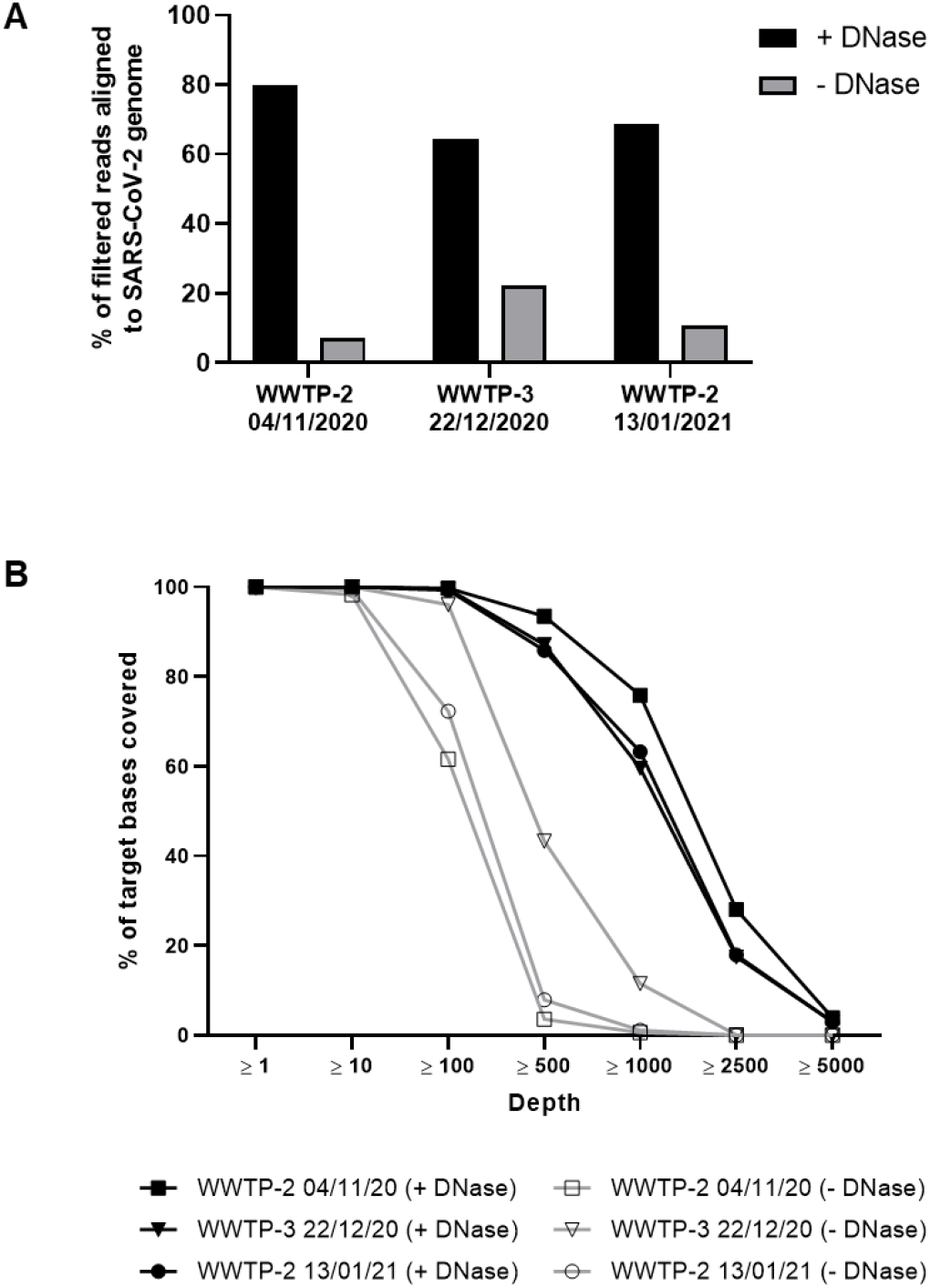
Analysis of wastewater nucleic acid for SARS-CoV-2 sequencing. Nucleic acid from the three wastewater collection sites and a pooled sample from the three collection sites at the indicated timepoints was used to generate SARS-CoV-2 amplicon libraries. Each dataset was randomly subsampled to 650,000 reads for comparison. (A) Percent of filtered reads aligned to SARS-CoV-2 genome for DNase-treated samples (+ DNase) or untreated total nucleic acid samples (- DNase). (B) Percent of target bases on the SARS-CoV-2 genome covered at or above indicated depths.

We aligned the reads a second time to a combined reference genome containing both SARS-CoV-2 and human reference genomes to examine the proportion of aligned, filtered reads attributable to contamination by human genetic material. Surprisingly, we found that the percentage of filtered reads aligning to the human genome increased after DNase treatment (3.5%-9.4% for total nucleic acid and 6.3%-16.5% for DNase-treated samples). However, the proportion of non-human contamination reduced by DNase treatment makes this the preferred protocol.

Finally, using all available reads, we analyzed the aligned sequencing reads from the DNase-treated samples to look for the presence of previously reported SARS-CoV-2 variants of concern associated with widespread viral strains. The variants that we detected (Supplemental file 1) were consistent with known variants in Dane County, Wisconsin during the sampling period^36^. Virtually all the SARS-CoV-2 sequences contained mutations corresponding to Nextstrain clade 20A. Mutations found in Nextstrain clades 20B, 20C, and 20G were also present, though mutations specific to strain 20B were present at 2.0% frequency in January, below the 10% frequency cutoff we had set for making a positive call. Although merely suggestive due to small sample size, this does correspond to Nextstrain reported data which also suggests a decrease in the prevalence of this strain over the sampling period. More transmissible strains were not observed in this data, consistent with Nextstrain reports for such strains initially appearing in this geographic area approximately 2 months after the sampling period.

## Conclusion

In this study, we describe a convenient, high throughput, robust, and consistent method to directly capture, concentrate, and detect total nucleic acids (TNA) from wastewater using silica based PureYield^®^ columns and optimized RT-qPCR. This method offers ease-of-use and minimizes the need for specialized laboratory equipment. In addition, the method achieves consistent recovery rates and significant reduction in RT-qPCR inhibitors.

During the course of this study, alternative direct capture methods for extraction of SARS-CoV-2 RNA from wastewater have been published, highlighting the simplicity of the general workflow^48^. Direct capture methods have also been applied in large-scale interlaboratory method assessment studies where they have shown superior and consistent performance over other methods including PEG/NaCl precipitation, centrifugal ultrafiltration, or charged membranes^49^. By using a protease treatment step, we are able to release a significant portion of viral nucleic acid associated with the solids in the wastewater. In addition, chaotropes and alcohols provide an effective nucleic acid binding environment for capture on a silica matrix. The two-step, modular purification strategy described in this work simplifies the workflow for users processing either small or large amounts of wastewater samples. The flexibility of the method and compatibility of the resulting nucleic acid with downstream analysis by RT-qPCR and SARS-CoV-2 sequencing allow for straightforward adoption for WBE-based viral surveillance approaches.

Throughout the early course of the COVID-19 pandemic, the ramifications of not having nationwide surveillance systems in place were observed. WBE and clinical diagnostic testing each can provide structured surveillance systems. WBE can complement clinical diagnostic testing by independently confirming prevalence of disease communities and possibly providing an early warning for future viral outbreaks. WBE also provides a low-cost tool to understand community spread in low resource areas. Similar to diagnostic clinical testing, WBE has experimental limitations (uncertainties related to timing and quantities of viral and viral nucleic acid shedding, RNA stability, effect of temperature, and sample processing techniques) that need to be well understood before using acquired data to inform epidemiological and public health efforts around the globe.

As the COVID-19 pandemic has progressed, SARS-CoV-2 genetic variants have arisen, often leading to increased transmissibility, concern about immune evasion, and subsequent outbreaks^50^. With the emergence of new strains, it will be informative to see how this rapidly evolving method is able to help understand the spread of variants within communities and guide health authorities to take appropriate measures^51–53^.

## Supporting information

medRxiv Supplementary Info

## Data Availability

Sequencing data are available upon request of the corresponding author.

## Data Statement

Sequencing data are available upon request of the corresponding author.

## Acknowledgements

Amplicon library preparation guidance, basic analysis guidance, and MiniSeq sequencing were provided by Swift Biosciences, Inc (https://swiftbiosci.com). We thank Paul Muschler for providing valuable feedback. We acknowledge the staff of Madison Metropolitan Sewage District (MMSD), Village of Oregon, WI and City of Sun Prairie, WI for their support and providing wastewater samples. We also thank the various research groups, public health, and commercial labs with whom we have had many productive discussions on this topic in these difficult times.

## Authorship contribution statement

**Subhanjan Mondal:** Conceptualization, Data curation, Formal analysis, Supervision, Methodology, Investigation, Roles/Writing - original draft, Writing - review & editing. **Nathan Feirer:** Conceptualization, Data curation Formal analysis, Methodology, Investigation, Roles/Writing - original draft, Writing - review & editing. **Michael Brockman:** Data curation, Formal analysis, Methodology, Roles/Writing - original draft, Writing - review & editing. **Melanie A. Preston:** Investigation, Writing - review & editing. **Sarah J. Teter:** Investigation, Writing - review & editing. **Dongping Ma:** Investigation, **Said A. Goueli:** Resources, Writing - review & editing. **Sameer Moorji**: Conceptualization, Resources. **Brigitta Saul:** Conceptualization, Resources. **James J. Cali:** Resources, Supervision, Writing - review & editing.

## References

1 Zhu, N. et al. A Novel Coronavirus from Patients with Pneumonia in China, 2019. N Engl J Med382, 727–733, doi:10.1056/NEJMoa2001017 (2020).

2 Hartenian, E. et al. The molecular virology of coronaviruses. J Biol Chem 295, 12910–12934, doi:10.1074/jbc.REV120.013930 (2020).

3 Cui, J., Li, F. & Shi, Z.-L. Origin and evolution of pathogenic coronaviruses. Nature Reviews Microbiology 17, 181–192, doi:10.1038/s41579-018-0118-9 (2019).

4 Zhou, P. et al. A pneumonia outbreak associated with a new coronavirus of probable bat origin. Nature 579, 270–273, doi:10.1038/s41586-020-2012-7 (2020).

5 He, X. et al. Temporal dynamics in viral shedding and transmissibility of COVID-19. Nat Med 26, 672–675, doi:10.1038/s41591-020-0869-5 (2020).

6 Cevik, M. et al. SARS-CoV-2, SARS-CoV, and MERS-CoV viral load dynamics, duration of viral shedding, and infectiousness: a systematic review and meta-analysis. The Lancet Microbe, doi:10.1016/S2666-5247(20)30172-5 (2020).

7 Furukawa, N., Brooks, J. & Sobel, J. Evidence Supporting Transmission of Severe Acute Respiratory Syndrome Coronavirus 2 While Presymptomatic or Asymptomatic. Emerging Infectious Disease journal 26, doi:10.3201/eid2607.201595 (2020).

8 Zhang, Z. et al. Early viral clearance and antibody kinetics of COVID-19 among asymptomatic carriers. 2020.2004.2028.20083139, doi:10.1101/2020.04.28.20083139 %J medRxiv (2020).

9 Park, S. K. et al. Detection of SARS-CoV-2 in Fecal Samples From Patients With Asymptomatic and Mild COVID-19 in Korea. Clin Gastroenterol Hepatol, doi:10.1016/j.cgh.2020.06.005 (2020).

10 Day, M. Covid-19: identifying and isolating asymptomatic people helped eliminate virus in Italian village. 368, m1165, doi:10.1136/bmj.m1165 %J BMJ (2020).

11 Wang, W. et al. Detection of SARS-CoV-2 in Different Types of Clinical Specimens. JAMA 323, 1843–1844, doi:10.1001/jama.2020.3786 %J JAMA (2020).

12 Chen, Y. et al. The presence of SARS-CoV-2 RNA in the feces of COVID-19 patients. J Med Virol 92, 833–840, doi:10.1002/jmv.25825 (2020).

13 Zheng, S. et al. Viral load dynamics and disease severity in patients infected with SARS-CoV-2 in Zhejiang province, China, January-March 2020: retrospective cohort study. BMJ 369, m1443, doi:10.1136/bmj.m1443 (2020).

14 Wolfel, R. et al. Virological assessment of hospitalized patients with COVID-2019. Nature 581, 465–469, doi:10.1038/s41586-020-2196-x (2020).

15 Han, M. S. et al. Viral RNA Load in Mildly Symptomatic and Asymptomatic Children with COVID-19, Seoul, South Korea. Emerging Infectious Disease journal 26, 2497, doi:10.3201/eid2610.202449 (2020).

16 Sethuraman, N., Jeremiah, S. S. & Ryo, A. Interpreting Diagnostic Tests for SARS-CoV-2. JAMA 323, 2249–2251, doi:10.1001/jama.2020.8259 (2020).

17 Lorenzo, M. & Picó, Y. Wastewater-based epidemiology: current status and future prospects. Current Opinion in Environmental Science & Health 9, 77–84, doi:https://doi.org/10.1016/j.coesh.2019.05.007 (2019).

18 Sims, N. & Kasprzyk-Hordern, B. Future perspectives of wastewater-based epidemiology: Monitoring infectious disease spread and resistance to the community level. Environ Int 139, 105689, doi:10.1016/j.envint.2020.105689 (2020).

19 Hellmer, M. et al. Detection of pathogenic viruses in sewage provided early warnings of hepatitis A virus and norovirus outbreaks. Appl Environ Microbiol 80, 6771–6781, doi:10.1128/AEM.01981-14 (2014).

20 Hovi, T. et al. Role of environmental poliovirus surveillance in global polio eradication and beyond. Epidemiol Infect 140, 1–13, doi:10.1017/S095026881000316X (2012).

21 Asghar, H. et al. Environmental surveillance for polioviruses in the Global Polio Eradication Initiative. J Infect Dis 210 Suppl 1, S294–303, doi:10.1093/infdis/jiu384 (2014).

22 Medema, G., Heijnen, L., Elsinga, G., Italiaander, R. & Brouwer, A. Presence o f SARS-Coronavirus-2 RNA in Sewage and Correlation with Reported COVID-19 Prevalence in the Early Stage of the Epidemic in The Netherlands. Environmental Science & Technology Letters, doi:10.1021/acs.estlett.0c00357 (2020).

23 La Rosa, G. et al. First detection of SARS-CoV-2 in untreated wastewaters in Italy. Sci Total Environ 736, 139652, doi:10.1016/j.scitotenv.2020.139652 (2020).

24 Ahmed, W. et al. First confirmed detection of SARS-CoV-2 in untreated wastewater in Australia: A proof of concept for the wastewater surveillance of COVID-19 in the community. Sci Total Environ 728, 138764, doi:10.1016/j.scitotenv.2020.138764 (2020).

25 Peccia, J. et al. Measurement of SARS-CoV-2 RNA in wastewater tracks community infection dynamics. Nature Biotechnology 38, 1164–1167, doi:10.1038/s41587-020-0684-z (2020).

26 Randazzo, W. et al. SARS-CoV-2 RNA in wastewater anticipated COVID-19 occurrence in a low prevalence area. Water Res 181, 115942, doi:10.1016/j.watres.2020.115942 (2020).

27 Wurtzer, S. et al. Evaluation of lockdown impact on SARS-CoV-2 dynamics through viral genome quantification in Paris wastewaters. 2020.2004.2012.20062679, doi:10.1101/2020.04.12.20062679 %J medRxiv (2020).

28 Bofill-Mas, S. & Rusiñol, M. Recent trends on methods for the concentration of viruses from water samples. Current Opinion in Environmental Science & Health 16, 7–13, doi:https://doi.org/10.1016/j.coesh.2020.01.006 (2020).

29 Ahmed, W. et al. Comparison of virus concentration methods for the RT-qPCR-based recovery of murine hepatitis virus, a surrogate for SARS-CoV-2 from untreated wastewater. Sci Total Environ 739, 139960, doi:10.1016/j.scitotenv.2020.139960 (2020).

30 Kitajima, M. et al. SARS-CoV-2 in wastewater: State of the knowledge and research needs. Science of The Total Environment 739, 139076, doi:https://doi.org/10.1016/j.scitotenv.2020.139076 (2020).

31 Pecson, B. M., Martin, L. V. & Kohn, T. Quantitative PCR for determining the infectivity of bacteriophage MS2 upon inactivation by heat, UV-B radiation, and singlet oxygen: advantages and limitations of an enzymatic treatment to reduce false-positive results. Appl Environ Microbiol 75, 5544–5554, doi:10.1128/AEM.00425-09 (2009).

32 Chen, S., Zhou, Y., Chen, Y. & Gu, J. fastp: an ultra-fast all-in-one FASTQ preprocessor. Bioinformatics 34, i884–i890, doi:10.1093/bioinformatics/bty560 %J Bioinformatics (2018).

33 Lander, E. S. et al. Initial sequencing and analysis of the human genome. Nature 409, 860–921, doi:10.1038/35057062 (2001).

34 Kent, W. J. et al. The Human Genome Browser at UCSC. 12, 996–1006, doi:10.1101/gr.229102 (2002).

35 McKenna, A. et al. The Genome Analysis Toolkit: A MapReduce framework for analyzing next-generation DNA sequencing data. 20, 1297–1303, doi:10.1101/gr.107524.110 (2010).

36 Hadfield, J. et al. Nextstrain: real-time tracking of pathogen evolution. Bioinformatics 34, 4121–4123, doi:10.1093/bioinformatics/bty407 (2018).

37 Karolchik, D. et al. The UCSC Table Browser data retrieval tool. Nucleic Acids Research 32, D493–D496, doi:10.1093/nar/gkh103 %J Nucleic Acids Research (2004).

38 Corman, V. M. et al. Detection of 2019 novel coronavirus (2019-nCoV) by real-time RT-PCR. Euro Surveill 25, doi:10.2807/1560-7917.ES.2020.25.3.2000045 (2020).

39 Colson, P. et al. Pepper Mild Mottle Virus, a Plant Virus Associated with Specific Immune Responses, Fever, Abdominal Pains, and Pruritus in Humans. PLOS ONE 5, e10041, doi:10.1371/journal.pone.0010041 (2010).

40 Kitajima, M., Sassi, H. P. & Torrey, J. R. Pepper mild mottle virus as a water quality indicator. npj Clean Water 1, 19, doi:10.1038/s41545-018-0019-5 (2018).

41 Bivins, A. et al. Cross-assembly phage and pepper mild mottle virus as viral water quality monitoring tools—potential, research gaps, and way forward. Current Opinion in Environmental Science & Health 16, 54–61, doi:https://doi.org/10.1016/j.coesh.2020.02.001 (2020).

42 Rosario, K., Symonds, E. M., Sinigalliano, C., Stewart, J. & Breitbart, M. <em>Pepper Mild Mottle Virus</em> as an Indicator of Fecal Pollution. 75, 7261–7267, doi:10.1128/AEM.00410-09 %J Applied and Environmental Microbiology (2009).

43 Schrader, C., Schielke, A., Ellerbroek, L. & Johne, R. PCR inhibitors – occurrence, properties and removal. 113, 1014–1026, doi:https://doi.org/10.1111/j.1365-2672.2012.05384.x (2012).

44 Bivins, A. et al. Persistence of SARS-CoV-2 in Water and Wastewater. Environmental Science & Technology Letters 7, 937–942, doi:10.1021/acs.estlett.0c00730 (2020).

45 Dada, A. C. & Gyawali, P. Quantitative microbial risk assessment (QMRA) of occupational exposure to SARS-CoV-2 in wastewater treatment plants. Science of The Total Environment, 142989, doi:https://doi.org/10.1016/j.scitotenv.2020.142989 (2020).

46 Fuqua, J. L. et al. A rapid assessment of wastewater for genomic surveillance of SARS-CoV-2 variants at sewershed scale in Louisville, KY. 2021.2003.2018.21253604, doi:10.1101/2021.03.18.21253604 %J medRxiv (2021).

47 Fontenele, R. S. et al. High-throughput sequencing of SARS-CoV-2 in wastewater provides insights into circulating variants. 2021.2001.2022.21250320, doi:10.1101/2021.01.22.21250320 %J medRxiv (2021).

48 Whitney, O. N. et al. Sewage, Salt, Silica, and SARS-CoV-2 (4S): An Economical Kit-Free Method for Direct Capture of SARS-CoV-2 RNA from Wastewater. Environ Sci Technol 55, 4880–4888, doi:10.1021/acs.est.0c08129 (2021).

49 Pecson, B. M. et al. Reproducibility and sensitivity of 36 methods to quantify the SARS-CoV-2 genetic signal in raw wastewater: findings from an interlaboratory methods evaluation in the U.S. Environmental Science: Water Research & Technology 7, 504–520, doi:10.1039/D0EW00946F (2021).

50 Mascola, J. R., Graham, B. S. & Fauci, A. S. SARS-CoV-2 Viral Variants —Tackling a Moving Target. JAMA 325, 1261–1262, doi:10.1001/jama.2021.2088 %J JAMA (2021).

51 Crits-Christoph, A. et al. Genome Sequencing of Sewage Detects Regionally Preval ent SARS-CoV-2 Variants. 12, e02703–02720, doi:10.1128/mBio.02703-20 %J mBio (2021).

52 Izquierdo-Lara, R. et al. Monitoring SARS-CoV-2 Circulation and Diversity through Community Wastewater Sequencing, the Netherlands and Belgium. Emerging Infectious Disease journal 27, 1405, doi:10.3201/eid2705.204410 (2021).

53 sLa Rosa, G. et al. Rapid screening for SARS-CoV-2 variants of concern in clinical and environmental samples using nested RT-PCR assays targeting key mutations of the spike protein. Water Research 197, 117104, doi:https://doi.org/10.1016/j.watres.2021.117104 (2021).

