## Supplementary material for "A Direct Capture Method for Purification and Detection of Viral Nucleic Acid Enables Epidemiological Surveillance of SARS-CoV-2": medRxiv Supplementary Info

Promega Corporation, 5430 E Cheryl Pkwy, Fitchburg, WI, 53711.

### Supplementary Information

| Oligo | Sequence | Reference |
|---|---|---|
| 2019-nCoV_N1-F | 5’-GACCCCAAAATCAGCGAAAT-3’ | US CDC |
| 2019-nCoV_N1-R | 5’-TCTGGTTACTGCCAGTTGAATCTG-3’ | US CDC |
| 2019-nCoV_N1-P | 5’-56-FAMACCCCGCAT-ZEN-TACGTTTGGTGGACC-IABkFQ-3’ | US CDC |
| 2019-nCoV_N2-F | 5’-TTACAAACATTGGCCGCAAA-IABkFQ-3’ | US CDC |
| 2019-nCoV_N2-R | 5’-GCGCGACATTCCGAAGAA-3’ | US CDC |
| 2019-nCoV_N2-P | 5’-56-FAM/ACAATTTGC-ZEN-CCCCAGCGCTTCAG-IABkFQ-3’ | US CDC |
| E_Sarbeco_F1 Fwd | 5’-ACAGGTACGTTAATAGTTAATAGCGT-3’ | Corman *et al.*[1] |
| E_Sarbeco_R2 Rev | 5’-ATATTGCAGCAGTACGCACACA-3’ | Corman *et al.*[1] |
| E_Sarbeco_P1 | 5’-56-FAM-ACACTAGCC-ZEN-ATCCTTACTGCGCTTCG-IABkFQ-3’ | Corman *et al.*[1] |
| PMMV FP | 5’-GAGTGGTTTGACCTTAACGTTGA-3’ | Kitajima *et al.*[2] |
| PMMV RP | 5’-TTGTCGGTTGCAATGCAAGT-3’ | Kitajima *et al.*[2] |
| PMMV P | 5’-Quasar 670-CCTACCGAAGCAAATG-BHQ1-3’ | Kitajima *et al.*[2] |
| MS2 F1 | 5’-TCCTAAAAGATGGAAACCCGATT-3’ | Zambenedetti *et al.*[3] |
| MS2 R1 | 5’-GGCCGGCGTCTATTAGTAGATG-3’ | Zambenedetti *et al.*[3] |
| MS2 P1 | 5’- FAM-CCTCAGCAATCGCAGCAAACTCCG-BHQ1-3’ | Zambenedetti *et al.*[3] |

| | | |
|---|---|---|
| CC169 | 5'-CATTCCGGATACTGCGATTTTAAGTG-3' | This Paper |
| NF001 | 5'-CGCCAAAAGCACTCTGATTGAC-3' | This Paper |
| NF002 | 5'-GCTTCCCCGACTTCTTTCGA-3' | This Paper |
| NF003 | 5'-CCAACCCTATTTTCATTCTTCGCC-3' | This Paper |
| NF004 | 5'-CTGGAAGATGGAAGCGTTTTGC-3' | This Paper |
| DS121-Org-560 | 5'- CAL Fluor Orange 560-CGCCCCCAGAAGCAATTTCGTGTAAA-BHQ1-3' | This Paper |

**Table S1**: Primers and probes used in this study. Oligos were ordered from either IDT (Coralville, IA) or Biosearch (Novato, CA)

| Pathogen | N1 | N2 | E |
|---|---|---|---|
| SARS-CoV-2 | + | + | + |
| OC43 | - | - | - |
| 229E | - | - | - |
| HKU1 | - | - | - |
| NL63 | - | - | - |
| Influenza A | - | - | - |
| RSV | - | - | - |
| *Legionella pneumophila* | - | - | - |
| *Pseudomonas aeruginosa* | - | - | - |

**Table S2**: The SARS-CoV-2 RT-qPCR detection kit described in the manuscript was tested for specificity of nucleic acid amplification using all three SARS-CoV-2 gene targets. Purified genomic DNA or RNA was purchased from commercial sources (American Type Culture Collection, ATCC) and amplification was performed using protocol described in the methods section. Plus sign indications presence of RT-qPCR signal from indicated pathogen.

### References

1 Corman, V. M. *et al.* Detection of 2019 novel coronavirus (2019-nCoV) by real-time RT-PCR. *Euro Surveill* **25**, doi:10.2807/1560-7917.ES.2020.25.3.2000045 (2020).

2 Kitajima, M., Sassi, H. P. & Torrey, J. R. Pepper mild mottle virus as a water quality indicator. *npj Clean Water* **1**, 19, doi:10.1038/s41545-018-0019-5 (2018).

3 Zambenedetti, M. R. *et al.* Internal control for real-time polymerase chain reaction based on MS2 bacteriophage for RNA viruses diagnostics. *Mem Inst Oswaldo Cruz* **112**, 339-347, doi:10.1590/0074-02760160380 (2017).
